# Prevalence of Antimicrobial Resistance in Tanzania: A Systematic Review and Meta-Analysis

**DOI:** 10.1101/2025.10.13.25337859

**Authors:** Charles B. Kafaiya, Johnson J. Mshiu, Obadia Bishoge, Jonathan M. Mshana, Sia Malekia, Irene Mremi, Angelina M. Lutambi, Mwanaada Kilima, Mary Mayige, Said Aboud

## Abstract

Antimicrobial resistance (AMR) threatens global health. Understanding resistance patterns aids in treatment and promotes responsible antimicrobial use. This review and meta-analysis assessed the prevalence of antimicrobial resistance among clinically relevant pathogens in Tanzania. A total of 18,265 studies identified from Google Scholar (18,000), PubMed (13), and Science Direct (252) underwent screening and full article review. Finally, 28 studies were included. A subgroup analysis was performed to evaluate the resistance patterns within antibiotic classes for specific pathogens. Descriptive statistics were used to describe the characteristics of the studies and the prevalence of antibiotic resistance. Heterogeneity was assessed using forest plots and the I² statistic. Among the included studies, most isolates (25.0%) were obtained from urine samples. Of these studies, 75% were cross-sectional studies and 92.9% were conducted in hospital settings. The analysis revealed high resistance to penicillin, particularly amoxicillin-clavulanic and ampicillin, with Klebsiella pneumoniae (0.96 [0.83–0.99]), Acinetobacter baumannii (0.94 [0.67–0.99]) and Escherichia coli (0.90 [0.81–0.95]). Similarly, erythromycin resistance was most prevalent in Campylobacter spp. (0.85 [0.80–0.89]). Ciprofloxacin resistance was highest in Acinetobacter baumannii (0.54 [0.33–0.73]), whereas amikacin resistance was highest in Proteus spp. (0.86 [0.35–0.99]). Ceftriaxone resistance was particularly high in Acinetobacter baumannii (0.91 [0.70–0.98]) and Pseudomonas aeruginosa (0.85 [0.74–0.92]). Resistance to meropenem was lowest among Escherichia coli (0.04 [0.01–0.10]) and Klebsiella spp. (0.07 [0.03–0.15]), with an overall pooled resistance to the ESKAPE-E pathogen of (0.11[0.06–0.19]). Imipenem and clindamycin each had an overall pooled resistance of (0.06[0.02-0.14]) against both Escherichia coli and Klebsiella pneumoniae. The findings highlight widespread resistance among key bacterial pathogens, ESKAPE-E, particularly in the *Access* and *Watch* groups of antibiotics. The variability in resistance patterns underscores the need to re-evaluate empirical treatment protocols (STG/NEMLIT) to ensure effective treatment regimens, strengthen antimicrobial stewardship, enhance surveillance systems, and promote rational antibiotic use.

## Introduction

Antimicrobial resistance occurs when pathogens resist exposure to antibiotics that are intended to kill or slow their growth (1,2). It has become one of the leading public health threats of the 21st century, posing significant dangers to global health, economic stability, and the effectiveness of healthcare systems (3). The World Health Organization (WHO) has recognised antimicrobial resistance as a serious public health concern that can result in prolonged hospital admissions, greater medical expenses and increased mortality rates (4). Furthermore, in 2017, the WHO identified ESKAPE pathogens; *Enterococcus faecium, Staphylococcus aureus, Klebsiella pneumoniae, Acinetobacter baumannii, Pseudomonas aeruginosa,* and *Enterobacter* species as major global health threats due to their rising antibiotic resistance and potential to cause serious infections in humans (2). This has made standard treatments less effective as bacterial pathogens change, making it more difficult to treat once-treatable infections. This phenomenon is particularly noticeable in low- and middle-income countries, where antibiotic abuse is pervasive, infectious disease burdens are high, and healthcare resources are scarce (3,5).

In Tanzania, the prevalence of antibiotic resistance has been increasingly documented; however, comprehensive data remain sparse (6,7). A variety of factors contribute to the rising rates of Antimicrobial Resistance (AMR) in the country, including over-prescription of antibiotics, self-medication, and inadequate regulatory frameworks for antibiotic use. Additionally, socio-economic factors such as personal awareness, attitudes, practices, poverty, lack of access to healthcare services, and poor sanitation and hygiene practices exacerbate the situation (8). The interplay of these factors creates an environment conducive to the emergence and spread of resistant strains of bacteria.

Despite the urgency of resistant pathogens, systematic reviews focusing specifically on Tanzania’s antibiotic resistance landscape are limited. While several studies have reported resistance patterns for specific pathogens, a consolidated analysis that captures the overall prevalence across the country is needed. This knowledge gap hampers effective policymaking and public health interventions aimed at curbing antibiotic misuse and managing resistance. By combining the available data, this systematic review and meta-analysis aimed to present a thorough overview of the prevalence of antibiotic resistance in Tanzania. A rigorous analysis of the published studies was conducted to identify the prevalence of resistant pathogens among different antibiotics. This review will inform the National Action Plan on Antimicrobial Resistance 2023 -2028 to strengthen surveillance, infection control, and antibiotic resistance stewardship programs in the country.

## Methodology

### Protocol registration

The systematic review and meta-analysis protocol was registered with PROSPERO under the ID number PROSPERO 2024 CRD42024608537.

### Settings

The evaluation was based on studies conducted in Tanzania, an East African country located in the African Great Lakes Region. It is bordered by Uganda to the northwest, Kenya to thee northeast, the Indian Ocean to the east, Mozambique and Malawi to the south, Zambia to the southwest, and Rwanda, Burundi, and the Democratic Republic of the Congo to the west. According to the 2022 national census, Tanzania has a population of approximately 62 million (9), making it the most populous country south of the equator. Antimicrobial resistance is a major public health issue in Tanzania, with a high prevalence of multidrug-resistant microorganisms (10). In clinical settings, the prevalence of multidrug-resistant bacteria varies between 25% and 50% (11). Previous research has shown a significant prevalence of methicillin-resistant *S. aureus* and extended-spectrum beta-lactamase-producing bacteria (7). The risk factors for antimicrobial resistance in Tanzania include irrational antibiotic use, inadequate hygiene and sanitation, transmission, sociodemographic characteristics, patient clinical information, and admission to healthcare facilities (12,13).

### Review procedures

The systematic review began by identifying the research question and was finalised with a report (14). The review adhered to the Preferred Reporting Items for Systematic Reviews and Meta-Analysis (PRISMA) guidelines for reviewing, analysing, and reporting systematic reviews (15), which covers all aspects of the manuscript, including the title, abstract, introduction, methods, results, and discussion.

### Search strategy

Relevant published studies were searched using electronic databases such as PubMed, ScienceDirect, and Google Scholar. The following phrases were used for the search: ((("Prevalence"[Mesh] OR "Prevalences" OR "Point Prevalence" OR "Point Prevalences" OR "Prevalence, Point" OR "Period Prevalence" OR "Period Prevalences" OR "Prevalence, Period") AND ("Risk Factors"[Mesh] OR "Factor, Risk" OR "Risk Factor" OR "Population at Risk" OR "Populations at Risk" OR "Risk Scores" OR "Risk Score" OR "Score, Risk" OR "Risk Factor Scores" OR "Risk Factor Score" OR "Score, Risk Factor" OR "Health Correlates" OR "Correlates, Health" OR "Social Risk Factors" OR "Factors, Social Risk" OR "Risk Factor, Social" OR "Risk Factors, Social" OR "Social Risk Factor")) AND ("Drug Resistance, Bacterial"[Mesh]OR "Antibacterial Drug Resistance" OR "Antibiotic Resistance, Bacterial")) AND ("Tanzania"[Mesh] OR "United Republic of Tanzania" OR "Zanzibar").

### Selection criteria and search outcomes

The following criteria were used to select relevant studies: (i) studies reporting on the prevalence of antibiotic resistance in bacterial pathogens; (ii) studies conducted in Tanzania over ten years, from 2014 to 2024; (iii) original cross-sectional studies, cohort studies, case-control studies, experimental studies, or modelling studies; and (iv) studies focused on antibiotic resistance. The review excluded studies that did not focus on antibiotic resistance, animal or laboratory-based studies without clinical relevance, reviews, letters, notes, editorials, and conference reports. This process yielded 18,265 articles related to the study topic. The studies were uploaded to Covidence, a web-based software (16) for screening, full article review, and data extraction. A total of 28 studies satisfied the set criteria and were included in this review (**Fig 1**).

**Fig 1.**
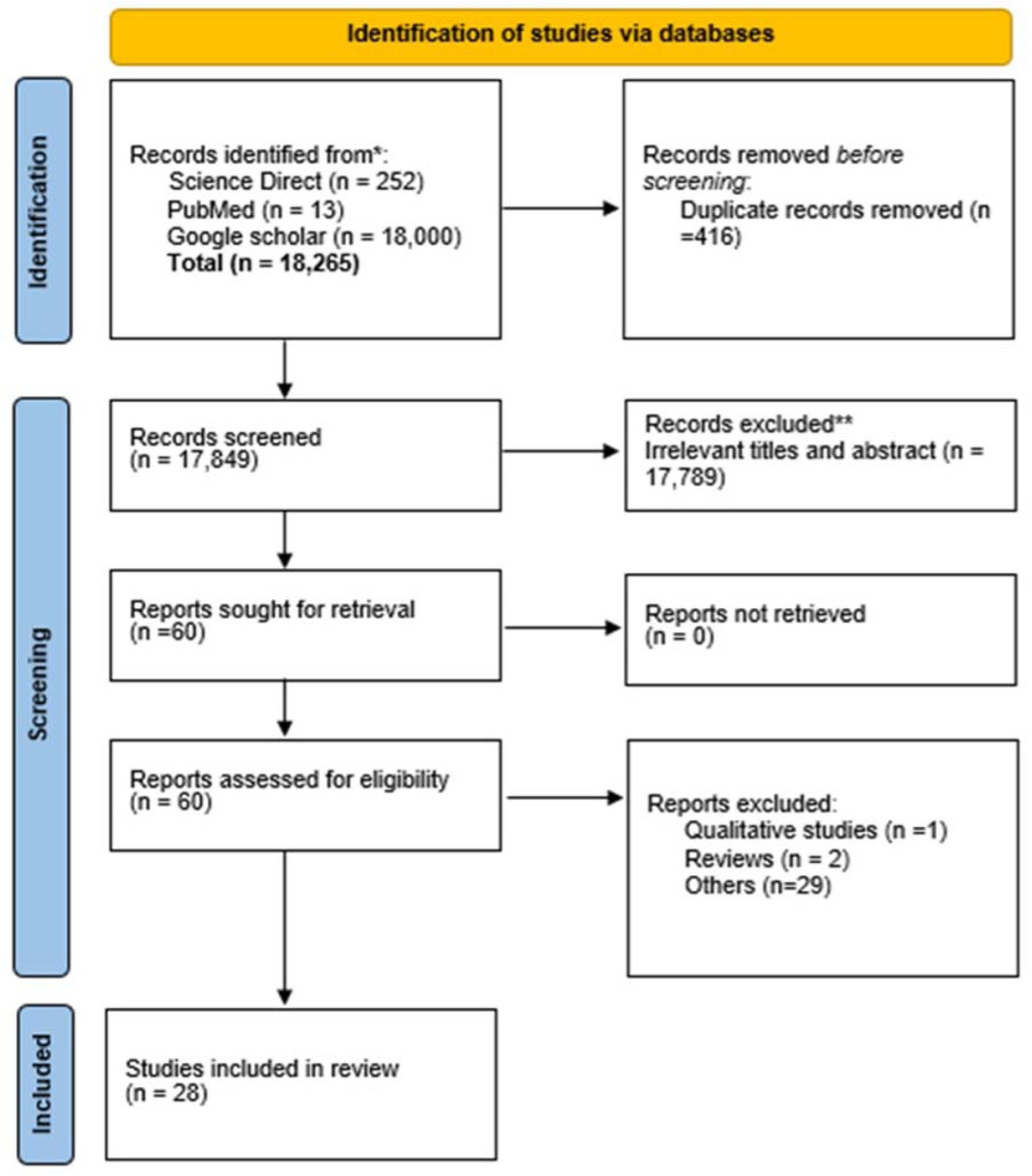
Prisma flow chart of the systematic review and article selection.

### Quality assessment

The included studies were evaluated for methodological quality and risk of bias using the Joanna Briggs Institute’s Quality Assessment Tool for Cross-Sectional Studies (17,18). The studies were scored as Yes, No, Unclear, and Not applicable (**S1 Table**).

### Data synthesis, analysis and reporting

The data were independently extracted by two researchers, focusing on various characteristics of the studies involved. These characteristics included the author and year of publication, study area, study design, study population, sampling methods, sample size, isolate samples, sources of samples, type of bacteria/pathogen, and type of antibiotic used. Descriptive statistics were used to summarise the characteristics of the studies using frequency and percentage. A meta-analysis was performed to estimate the pooled prevalence rates of antibiotic resistance using random-effects models. Forest plots and I² statistics were used to evaluate heterogeneity among the studies, with higher values indicating significant heterogeneity (19). Subgroup analyses were conducted to investigate potential drug resistance in specific pathogens associated with each antibiotic. All analyses were conducted using the R libraries (meta) and (metafor) (20,21). Finally, the results were visually presented through tables and figures, including bar charts and plot forests.

## Results

### Study characteristics

#### Settings, study designs and year of publication of the selected studies

Of the 28 included studies, most (32%) were conducted in Dar es Salaam, followed by Mwanza (25%) and Kilimanjaro (17.9%), with 75.0% being cross-sectional. Additionally, 17.9% of the studies were published in 2020, while 14.3% were published in 2016, 2019, and 2022 (**Fig 2**).

**Fig 2.**
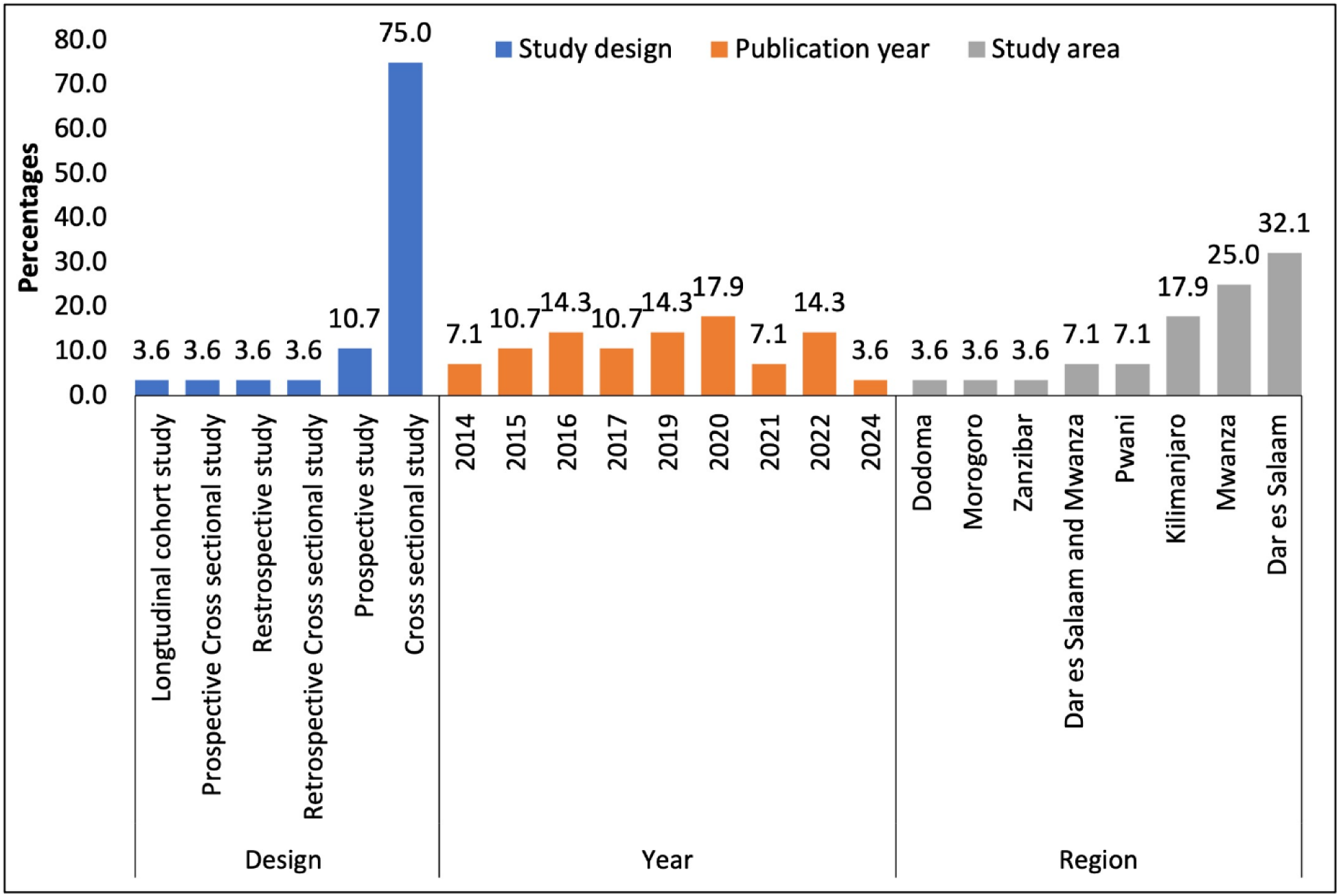
Distribution of the Year of Publication, Study Design, and Region.

#### Study population, sample size and number of isolates

Most of the selected studies (46%) involved children under five years of age. The sample size of the selected studies ranged from 75 (22) to 4,306 (23), with the number of isolates ranging from 22 (24) to 4,030 (23) (**Table 1**). The most common clinical samples were urine (25%), nasopharyngeal swabs (10.7%), open wound pus swabs (7.1%), and blood (3.6%) (**Fig 3**).

**Fig 3.**
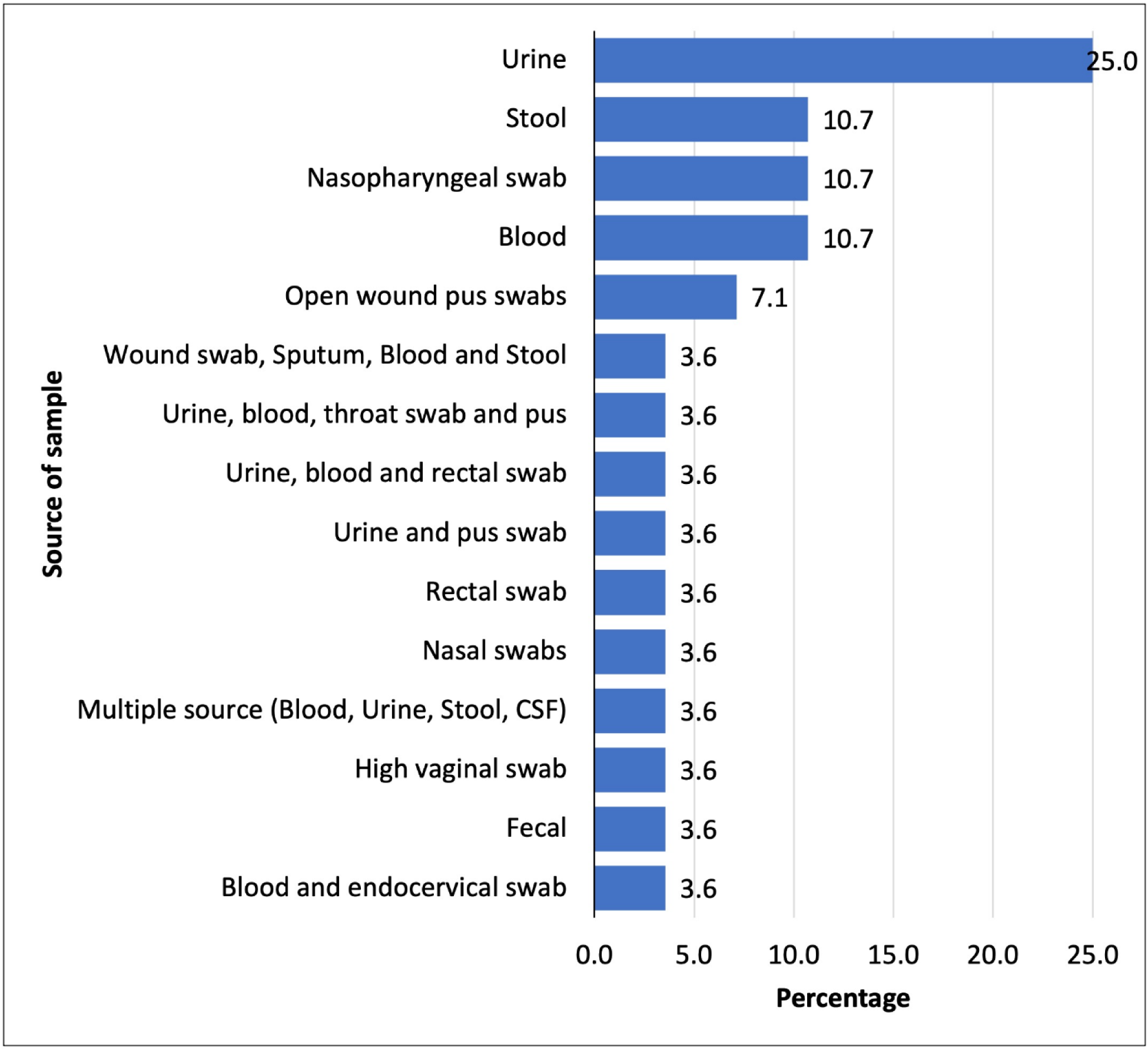
Sources of samples for selected studies.

**Table 1:** Study population, sampling methods, sample size, number of isolates and sample sources of the selected studies.

| **Author** | **Study population** | **Sample Size** | **Number of Isolates** |
| --- | --- | --- | --- |
| Manyahi et al., 2014 (25) | Patients with clinical evidence of SSI | 100 | 147 |
| Moyo et al., 2014 (26) | Healthy under-fives | 114 | 114 |
| Marwa et al., 2015 (27) | HIV/AIDS patients | 945 | 155 |
| Komba et al., 2015 (28) | Patients with enteric and non-enteric symptoms | 1195 | 136 |
| Ahmed et al., 2015 (29) | Malnourished under-fives | 402 | 84 |
| Okamo et al., 2016 (30) | Medical students | 314 | 66 |
| Moremi et al., 2016 (31) | More samples were from paediatric population | 3330 | 439 |
| Tellevik et al., 2016 (32) | Children Below 2 years | 603 | 284 |
| Seidman et al., 2016 (33) | Young children | 377 | 2492 |
| Ahmed et al., 2017 (34) | Malnourished under-fives | 402 | 42 |
| Joachim et al., 2017 (24) | Patients attended at EMD/IPD | 258 | 22 |
| Kumburu et al., 2017 (35) | Patient admitted in surgical and medical department | 575 | 377 |
| Kiponza et al., 2019 (36) | Post-delivery women | 197 | 107 |
| Seni et al., 2019 (37) | Pregnant women with significant bacteriuria | 1828 | 323 |
| Emgård et al., 2019 (38) | Children | 775 | 244 |
| Madut et al., 2019 (39) | Adolescent and adult patients aged $\geq 13$ y with fever. | 1648 | 31 |
| Daud et al., 2020 (40) | Children | 282 | 282 |
| Kamgobea et al., 2020 (41) | Pregnant women with or without PROM | 350 | 39 |
| Mikomangwa et al., 2020 (42) | Patients attended | 183 | 201 |
| Manyahi et al., 2020 (43) | All inpatients with features suggestive of BSI | 402 | 44 |
| Kadigi et al., 2020 (44) | Under-fives with symptoms of UTI | 270 | 80 |
| Letara et al., 2021 (45) | Children | 350 | 76 |
| Sangeda et al., 2021 (46) | Under-fives with symptoms of UTI | 214 | 35 |
| Manyahi et al., 2022 (47) | Admitted patients | 198 | 179 |
| Masoud et al., 2022 (22) | Admitted patients | 75 | 76 |
| Schmider et al., 2022 (48) | Outpatients aged $\geq 2$ years with symptoms of UTI | 270 | 119 |
| Silago et al., 2022 (49) | Patients who resided within the study area | 1327 | 412 |
| Muhiddin Hamada Omar<br>2024 (23) | Hospitalized patients | 4306 | 4030 |

### Results from meta-analysis

The meta-analysis included 28 studies that investigated antibiotic resistance patterns across a broad spectrum of pathogens, including ESKAPE-E bacteria (*Escherichia coli, Staphylococcus aureus, Klebsiella pneumoniae, Acinetobacter baumannii, Pseudomonas aeruginosa,* and *Enterobacter spp.*), *Proteus spp., Streptococcus spp., Campylobacter spp.,* and *P. mirabilis*.

The review examined various pathogens against 17 antibiotics, including penicillin (amoxicillin and ampicillin), macrolides (erythromycin), fluoroquinolones (ciprofloxacin), aminoglycosides (amikacin), tetracyclines (tetracycline), sulphonamides (trimethoprim-sulfamethoxazole), and cephalosporins (2^nd^,3^rd^ and 4^th^ generations). It also included carbapenems (meropenem and imipenem), lincosamides (clindamycin), and nitrofuran compounds (nitrofurantoin) (**S1-S14 Figs) (Fig. 4-6**).

**Fig 4.**
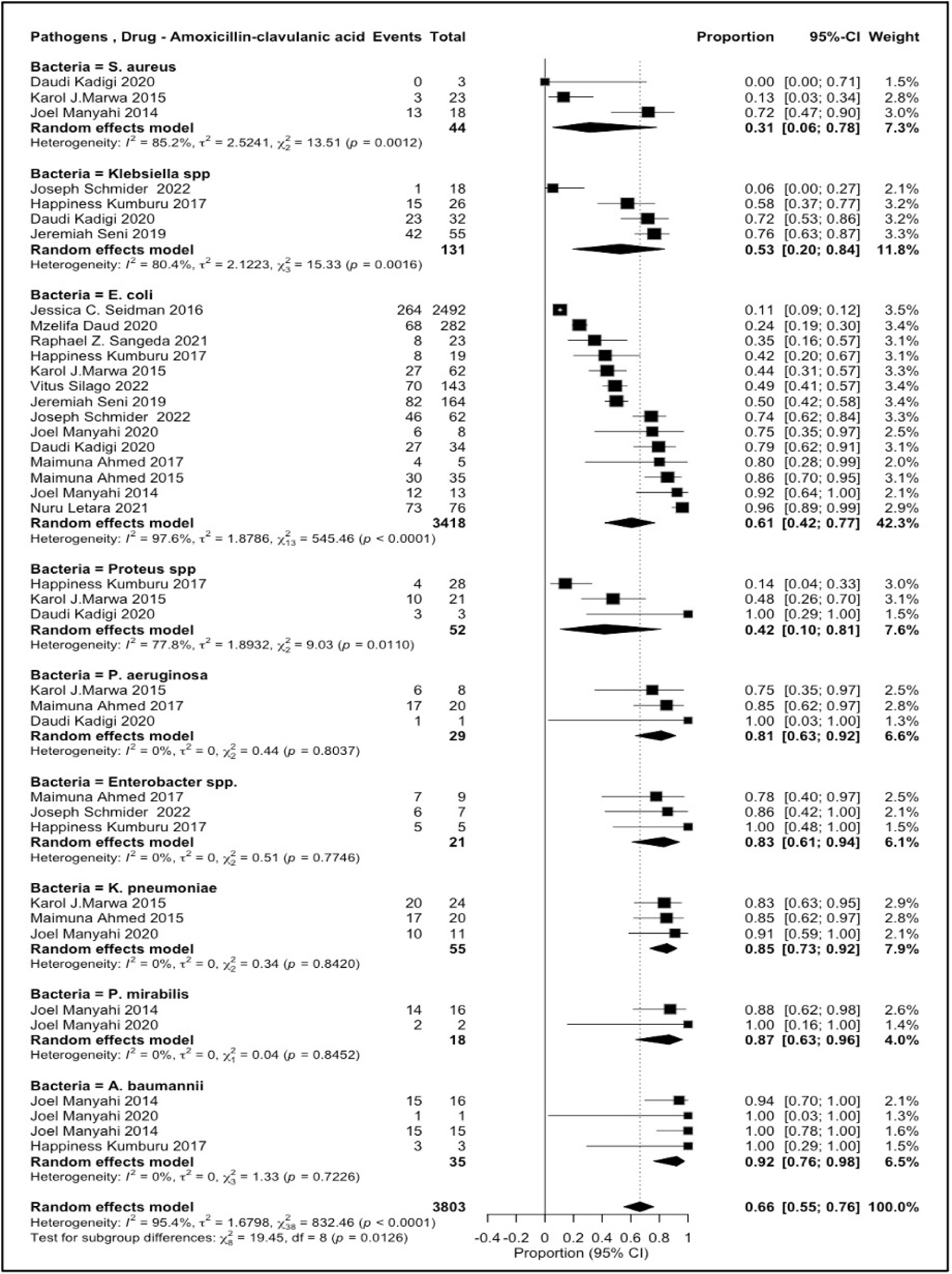
Amoxicillin/clavulanic acid resistance patterns among various pathogens.

**Fig 5.**
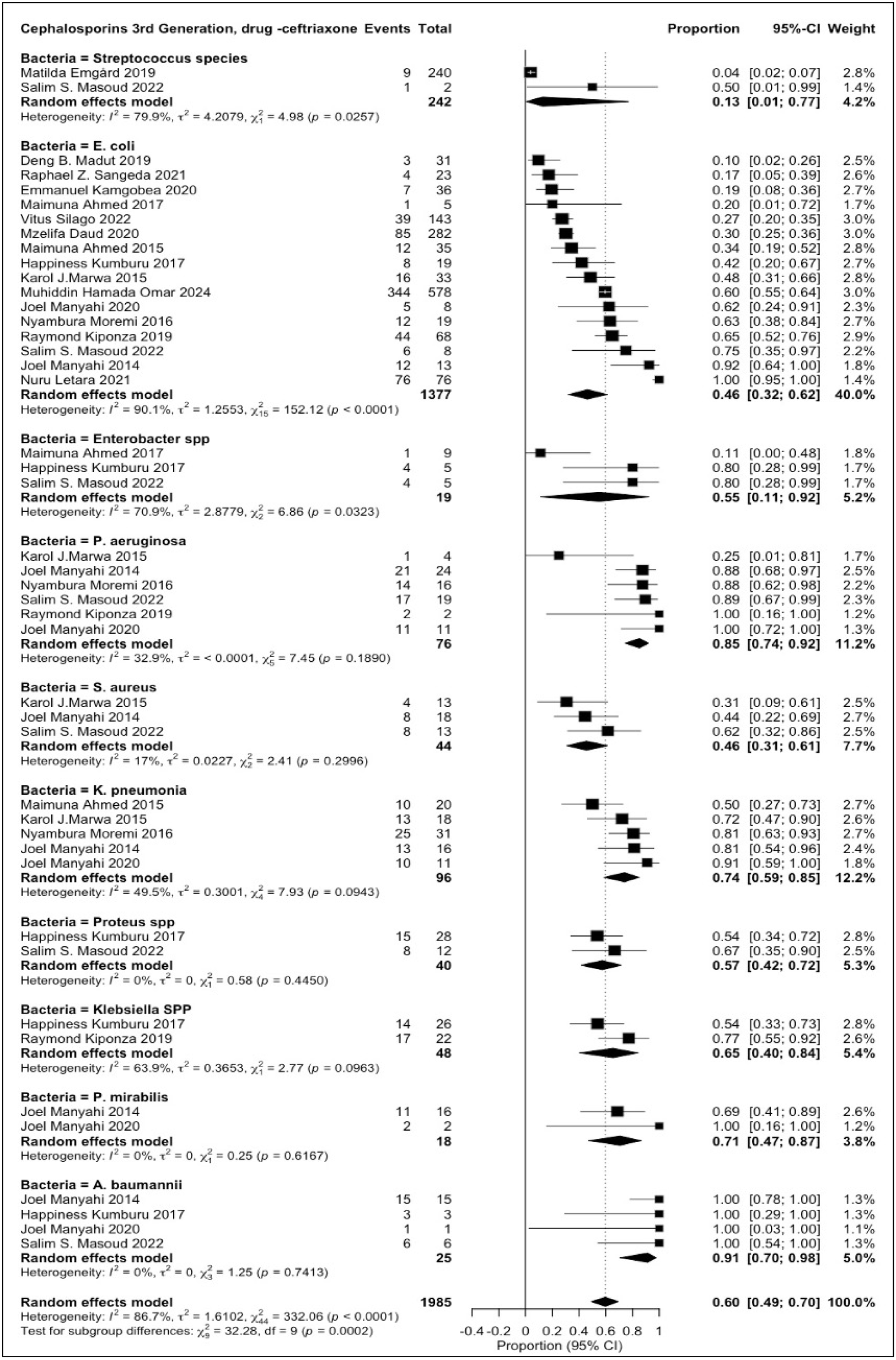
Ceftriaxone resistance patterns in various pathogens.

**Fig 6.**
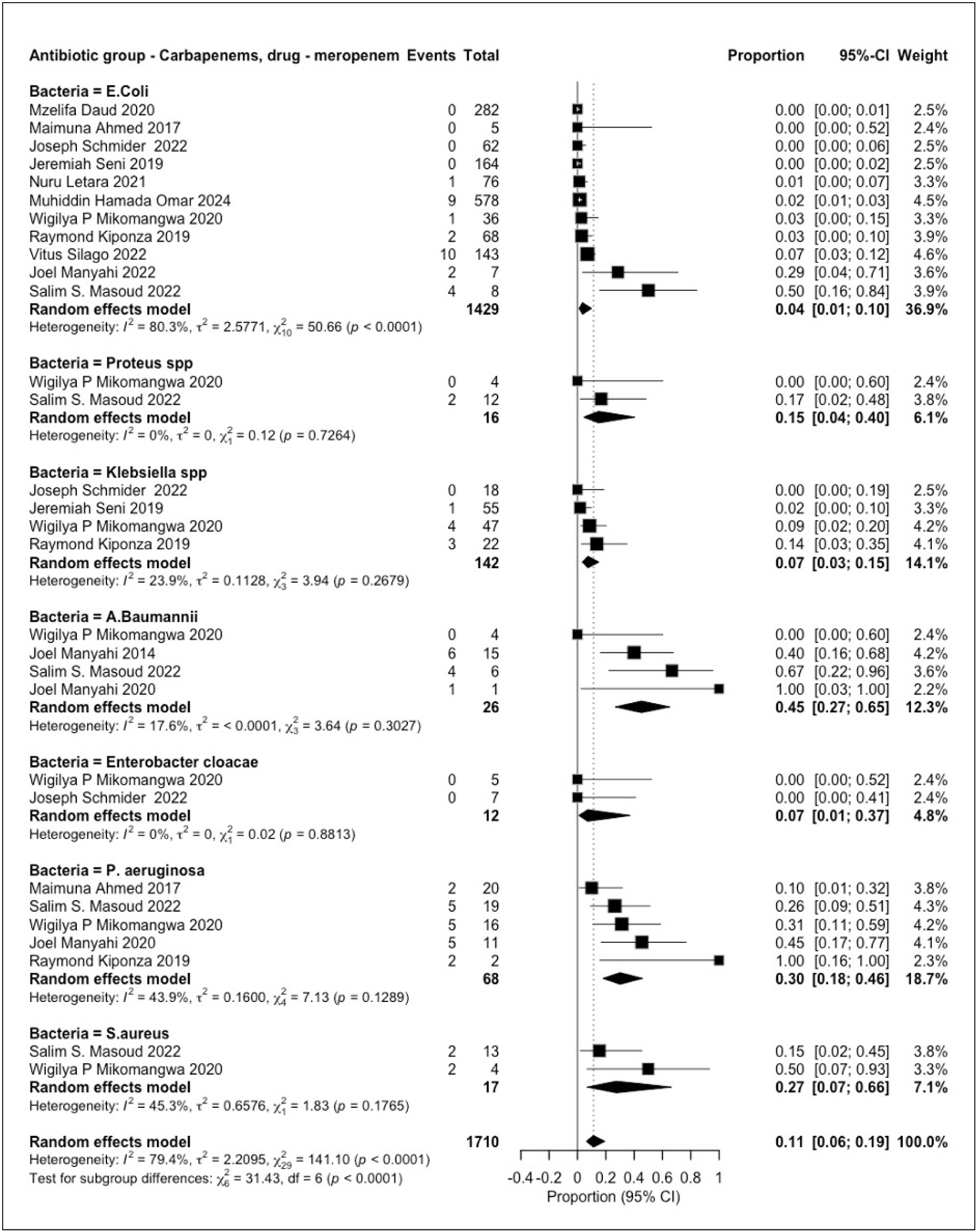
Meropenem resistance patterns among various pathogens.

A subgroup analysis was conducted to assess the resistance patterns within each antibiotic class for specific pathogens. The results revealed high resistance to penicillin (*amoxicillin–clavulanic acid*), with the highest pooled prevalence observed in *Acinetobacter baumannii* (0.92 [0.76–0.98]), followed by *Proteus mirabilis* (0.87 [0.63–0.96]), *Klebsiella pneumoniae* (0.85 [0.73–0.92]), and *Enterobacter spp.* (0.83 [0.61–0.94]). Resistance was also notable among *Escherichia coli* (0.61 [0.42–0.77]) and *Klebsiella spp.* (0.53 [0.20–0.84]) (**Fig 1**). Similarly, a high level of resistance to penicillin (ampicillin) was observed among ESKAPE- E pathogens and *Proteus* spp. The pooled prevalence of resistance ranged from *Klebsiella pneumoniae* (0.96 [0.83–0.99]), *Acinetobacter baumannii* (0.94 [0.67–0.99]), *Escherichia coli* (0.90 [0.81–0.95]), *Klebsiella* spp. (0.90 [0.81–0.95]), *Enterobacter cloacae* (0.78 [0.46–0.94]), *Pseudomonas aeruginosa*(0.77 [0.47–0.92]), *Proteus*spp. (0.76 [0.58–0.88]), and *Staphylococcus aureus* (0.76 [0.38–0.94]) (**S1 Fig**).

Resistance to macrolides, specifically erythromycin, was highest among *Campylobacter spp.* (0.85 [0.80–0.89]), followed by *Escherichia coli* (0.61 [0.16–0.93]) and *Staphylococcus aureus* (0.52 [0.35–0.68]) (**S2 Fig**). Ciprofloxacin exhibited a pooled resistance prevalence of 0.54 [0.33–0.73] in *Acinetobacter baumannii* (**S3 Fig**), whereas amikacin resistance was notably high in *Proteus spp.* (0.86 [0.35–0.99]) (**S4 Fig**). Tetracycline resistance was prevalent in *Klebsiella pneumoniae* (0.81 [0.24–0.98]) and *Escherichia coli* (0.77 [0.71–0.83]) (**S5 Fig**). Additionally, trimethoprim-sulfamethoxazole showed high resistance in *Klebsiella pneumoniae* (0.93 [0.77–0.98]), *Escherichia coli* (0.86 [0.75–0.93]), and *Proteus mirabilis* (0.80 [0.30–0.97]) (**S6 Fig**).

The pathogens exhibiting the highest resistance to ceftriaxone, a third-generation cephalosporin, include *Acinetobacter baumannii* (0.91 [0.70–0.98]), *Proteus mirabilis* (0.71 [0.47–0.87]), *Klebsiella species* (0.65 [0.40–0.84]), *Klebsiella pneumoniae* (0.74 [0.59–0.85]), and *Pseudomonas aeruginosa* (0.85 [0.74–0.92]) (**Fig 2**). Similarly, high resistance to cefotaxime was observed in *Acinetobacter baumannii* (0.92 [0.53–0.99]), *Pseudomonas aeruginosa* (0.90 [0.53–0.99]), *Klebsiella pneumoniae* (0.94 [0.62–0.99]), *Escherichia coli* (0.81 [0.35–0.97]), and *Proteus mirabilis* (0.81 [0.24–0.98]) (**S7 Fig**).

For ceftazidime, the highest resistance was noted among *Klebsiella pneumoniae* (0.89 [0.64–0.97]), *Acinetobacter baumannii* (0.82 [0.58–0.93]), and *Proteus mirabilis* (0.67 [0.44– 0.84]) (**S8 Fig**). Resistance to the fourth-generation cephalosporin, cefepime, was also observed across multiple bacterial species, with *Enterobacter species* (0.91 [0.66–0.98]), *Escherichia coli* (0.86 [0.14–1.00]), *Klebsiella species* (0.73 [0.59–0.83]), and *Citrobacter species* (0.72 [0.47– 0.88]) ( **S9 Fig**).

Despite the overall high resistance observed among pathogens against specific antibiotics, certain pathogens exhibited a lower pooled resistance prevalence. *Escherichia coli* and *Staphylococcus aureus* showed resistance to cefuroxime (0.38 [0.19–0.60]) and cefoxitin (0.38 [0.26–0.53]), respectively (**S10**–**S11 Figs**). The ESKAPE-E pathogens were tested against meropenem, exhibiting an overall pooled resistance prevalence of 0.11 [0.06–0.19]). Among individual pathogens, *Escherichia coli* showed the lowest resistance (0.04 [0.01–0.10]), followed by *Klebsiella species* (0.07 [0.03–0.15]), *Enterobacter cloacae* (0.07 [0.01–0.37]), *Pseudomonas aeruginosa* (0.30 [0.18–0.46] ), and *Staphylococcus aureus* (0.27 [0.07–0.66]) (**Fig 3**). Notably, *Acinetobacter baumannii* exhibited the highest resistance (0.45 [0.27–0.65] ). For imipenem, *Klebsiella pneumoniae* and *Escherichia coli* had an overall pooled resistance prevalence of 0.06 [0.02–0.26], with individual resistance rates of (0.08 [0.02–0.26]) and (0.06 [0.02–0.26]), respectively (**S12 Fig)**. Similarly, *Klebsiella pneumoniae* and *Escherichia coli* exhibited an overall pooled resistance prevalence of (0.06 [0.02–0.14]) to clindamycin (**S13 Fig**).

In the case of nitrofurantoin, *Staphylococcus aureus*, *Escherichia coli*, and *Klebsiella species* demonstrated an overall pooled resistance prevalence of (0.24 [0.16–0.34]) (**S14 Fig**). The included studies demonstrated moderate to substantial heterogeneity across pathogens and antibiotics, with 76.5% exhibiting high heterogeneity (above 75%).

## Discussion

This systematic review investigated the prevalence of antimicrobial resistance in Tanzania. The findings indicate alarmingly high resistance rates to the *AWaRe (Access, Watch, Reserve)* classes of antibiotics in Tanzania. The ESKAPE-E pathogens have exhibited high resistance to a wide range of these groups of antibiotics, which suggests a significant challenge in treating infections caused by these pathogens in the country (2).

ESKAPE-E pathogens are the leading causes of urinary tract infections, surgical site infections, skin infections, upper and lower respiratory infections, bloodstream infections, and nosocomial infections in Tanzania (10, 50). The current study identified high resistance rates among these pathogens to commonly used *Access group* of antibiotics, including *nitrofurantoin (24%), erythromycin (56%), amoxicillin-clavulanic acid (66% ), Tetracycline( 67%), and ampicillin (86%).* A study conducted in South Africa and a review conducted in East Africa from 2005 to 2015, involving Uganda, Kenya, Ethiopia, Rwanda, and the Democratic Republic of Congo, reported similar resistance patterns(51,52). These elevated resistance levels significantly compromise treatment efficacy, leading to prolonged illness, increased healthcare costs, and higher mortality rates (6,53).

Considering individual pathogens and specific *Access group* of antibiotics, resistance to *ampicillin* was particularly alarming, with 96% of *Klebsiella pneumoniae*, 94% of *Acinetobacter baumannii*, and 90% of *Escherichia coli* isolates exhibiting resistance. Given that ampicillin is recommended as a treatment option for these pathogens in the Tanzania Standard Treatment Guidelines (54), such high resistance raises concerns about treatment complications and prolonged hospital stays. These findings align with previous studies conducted in South Africa and East African countries (51,52), further emphasising the need for urgent interventions.

Moreover, *Acinetobacter baumannii*, *Proteus mirabilis*, and *Klebsiella pneumoniae* exhibited high resistance rates of 92 %, 87 %, and 85 %, respectively, to *amoxicillin-clavulanic acid*, complicating the effective management of bloodstream infections, wound infections, urinary tract infections (UTIs), upper respiratory infections, and pneumonia(25,50,55–56). Similar resistance patterns have been reported in previous studies(57), highlighting the growing challenge of antimicrobial resistance in those pathogens. However, nitrofurantoin, commonly recommended for UTIs treatment in Tanzania (54), showed relatively lower resistance rates, with *Escherichia coli* (21%), *S. aureus* (24%), and *Klebsiella spp.* (32%). These findings are consistent with those of other studies (52), suggesting that nitrofurantoin remains a viable treatment option for UTIs, although continuous monitoring is essential.

Furthermore, more than half of the samples tested for *erythromycin* susceptibility exhibited resistance, to *Campylobacter spp.* (85%), *Escherichia coli* (61%), and *Staphylococcus aureus* (52%) showed notable resistance levels. This trend aligns with the findings of studies conducted in Kenya (58) and a review conducted in Africa (59), reinforcing concerns regarding the declining effectiveness of erythromycin.

Similarly, the *Watch group* of antibiotics revealed an overall resistance to *sulfonamides (84%)*, *ceftriaxone*(60%), *ceftazidime(55%)*, *cefuroxime(38%),* and *ciprofloxacin(34%).* Ceftriaxone, a crucial antibiotic for severe bloodstream infections, surgical site infections, complicated UTIs, and gastrointestinal infections, has shown higher resistance to a wide range of pathogens. *A. baumannii* demonstrated a resistance of 91%, followed by *Pseudomonas aeruginosa* (85%) and *K. pneumoniae* (74%). These findings are consistent with a review conducted focusing on *A. baumannii* in 2019 (60), which reported similar high resistance levels, further emphasising the limited efficacy of ceftriaxone in treating severe bacterial infections.

Moreover, Ciprofloxacin resistance was assessed among ESKAPE-E pathogens, with more than half of *A. baumannii* demonstrating resistance, whereas other pathogens exhibited resistance rates below 50%. This high resistance in A. *baumannii* may indicate selective pressure due to the widespread use of ciprofloxacin, particularly in the management of urinary tract infections (UTIs), post-operative care, and bloodstream infections (54,61–62). Despite the observed resistance, ciprofloxacin remains effective in treating these infections. However, caution is required to prevent further resistance development through prudent use and antimicrobial stewardship programs.

The *reserved group* of antibiotics, intended for last-resort use, including *carbapenems, amikacin*, *clindamycin,* and *cefepime*, revealed heterogeneous resistance ranging from as low as 6% to as high as 81%. This trend raises concerns about treatment outcomes for complicated cases referred from lower-level facilities, where *Access to group* antibiotics has failed. When analysing resistance by specific antibiotic-pathogen combinations, both *clindamycin* and *imipenem* exhibited relatively low resistance rates against *E. coli* (8%) and *K. pneumoniae* (3%), respectively. The lower resistance observed for these antibiotics can be attributed to restricted access, as they are not available over the counter, thereby reducing misuse and selective pressure (63). These findings align with studies conducted in Uganda and South Africa(64,65), which also reported lower resistance rates to these reserved antibiotics.

Among ESKAPE-E pathogens, *meropenem* demonstrated relatively low resistance, with *E. coli* (4%), *Klebsiella spp.* (7%), and *Enterobacter cloacae* (7%) showing low resistance.

However, *A. baumannii* showed notably higher resistance, which may reflect its well-documented ability to develop multidrug resistance (60). These findings underscore the importance of maintaining strict control over the use of *reserved* antibiotics while implementing continuous surveillance to prevent resistance escalation. Similar trends have been observed in studies conducted in South Africa and Ethiopia (64,66,67), further supporting the need for continuous monitoring. Conversely, *cefepime* and *amikacin* exhibited significantly higher resistance rates, particularly in *E. coli* (86% and 20%, respectively). Alarmingly, elevated resistance was also observed in *Enterobacter spp.* (91% resistant to *cefepime* ) and *Proteus spp.* (86% resistant to *amikacin*), suggesting that these previously reserved antibiotics may be losing their efficacy. These findings are similar to those of studies conducted in Iran, USA, India, and Africa (68–71), where higher resistance rates were reported, highlighting the potential regional differences in antibiotic use and resistance patterns.

This review provides valuable insights into the pooled prevalence of antimicrobial resistance in Tanzania. However, this study has some limitations. Most studies were conducted in hospital settings, which may limit the generalisability of the findings to the broader population. Similarly, over half of the studies were conducted in urban areas, particularly in Dar es Salaam and Mwanza.

## Conclusion

The current review highlights the critical and troubling rates of antimicrobial resistance among *ESKAPE-E* pathogens in Tanzania, which create significant challenges in the effective management of urinary tract infections (UTIs), upper respiratory tract, skin, bloodstream, pneumonia, and wound infections. The high resistance to the *Access* and *Watch* groups of antibiotics indicates that standard treatment regimens may no longer be reliable, resulting in longer illnesses, increased healthcare expenses, and higher mortality rates. Although resistance levels to reserved antibiotics like *carbapenems, cefepime* and *amikacin* are relatively low, troubling trends have been observed, especially with *A. baumannii, E. coli, Proteus spp* and *Enterobacter spp*. This situation emphasises the need to protect these last-resort treatments through rigorous antimicrobial stewardship and regulatory policies, and by revising the Tanzania Standard Treatment Guidelines. Moreover, implementing prescription regulations is essential for combating the ongoing development of antimicrobial resistance.

## Supporting information

Supplementary Figures_Results

PRISMA Checklist

## Data Availability

All data produced in the present study are available upon reasonable request to the authors

## Abbreviations

ESKAPE-E: *Enterococcus faecium, Staphylococcus aureus, Klebsiella pneumoniae, Acinetobacter baumannii, Pseudomonas aeruginosa and Enterobacter* spp, *Escherichia coli*
STG: Standard Treatment Guideline
NEMLIT: The National Essential Medicines List of Tanzania
AMR: Antimicrobial resistance
BSI: bloodstream infections)
SSI: Surgical Site Infection)
*AWaRe*: Access, Watch, Reserve
*Access*: Penicillin, Erythromycin, Tetracycline, Nitrofurantoin
*Watch*: Cephalosporins, Ciprofloxacin, Sulphonamides
*Reserve*: Carbapenems, Cefepime, Clindamycin, Amikacin

## Acknowledgement

The authors would like to thank the researchers who published their articles online, without which this review would not have been possible.

## Authors contribution

CBK, OB and JM (conceptualization, protocol registration, literature search, full article review, writing the manuscript), JJM (conceptualization, data analysis and interpretation), SM and IM (Screening and data extraction), AML (Screening, data extraction and review) and MK, MM and SA (review of the manuscript)

## Data availability

The datasets used in this review are available upon request from the corresponding author. All relevant data are provided separately as supplementary files.

## Supporting information

**S1 Table**. Quality Assessment Table

**S1 Fig.** Ampicillin resistance patterns among various pathogens

**S2 Fig.** Erythromycin resistance patterns among various pathogens

**S3 Fig.** Ciprofloxacin resistance patterns among various pathogens

**S4 Fig.** Amikacin resistance patterns among various pathogens

**S5 Fig.** Tetracycline resistance patterns among various pathogens

**S6 Fig.** Trimethoprim-sulphamethoxazole resistance patterns among various pathogens

**S7 Fig.** Cefotaxime resistance patterns among various pathogens

**S8 Fig.** Ceftazidime resistance patterns among various pathogens

**S9 Fig.** Cefepime resistance patterns among various pathogens

**S10 Fig.** Cefuroxime resistance patterns with E.coli

**S11 Fig.** Cefoxitin resistance patterns among various pathogens

**S12 Fig.** Imipenem resistance patterns among various pathogens

**S13 Fig.** Clindamycin resistance patterns among various pathogens

**S14 Fig.** Nitrofurantoin resistance patterns among various pathogens

**S1 Fig A1**. PRISMA 2020 Checklist

