## Supplementary Figures_Results for "Prevalence of Antimicrobial Resistance in Tanzania: A Systematic Review and Meta-Analysis"

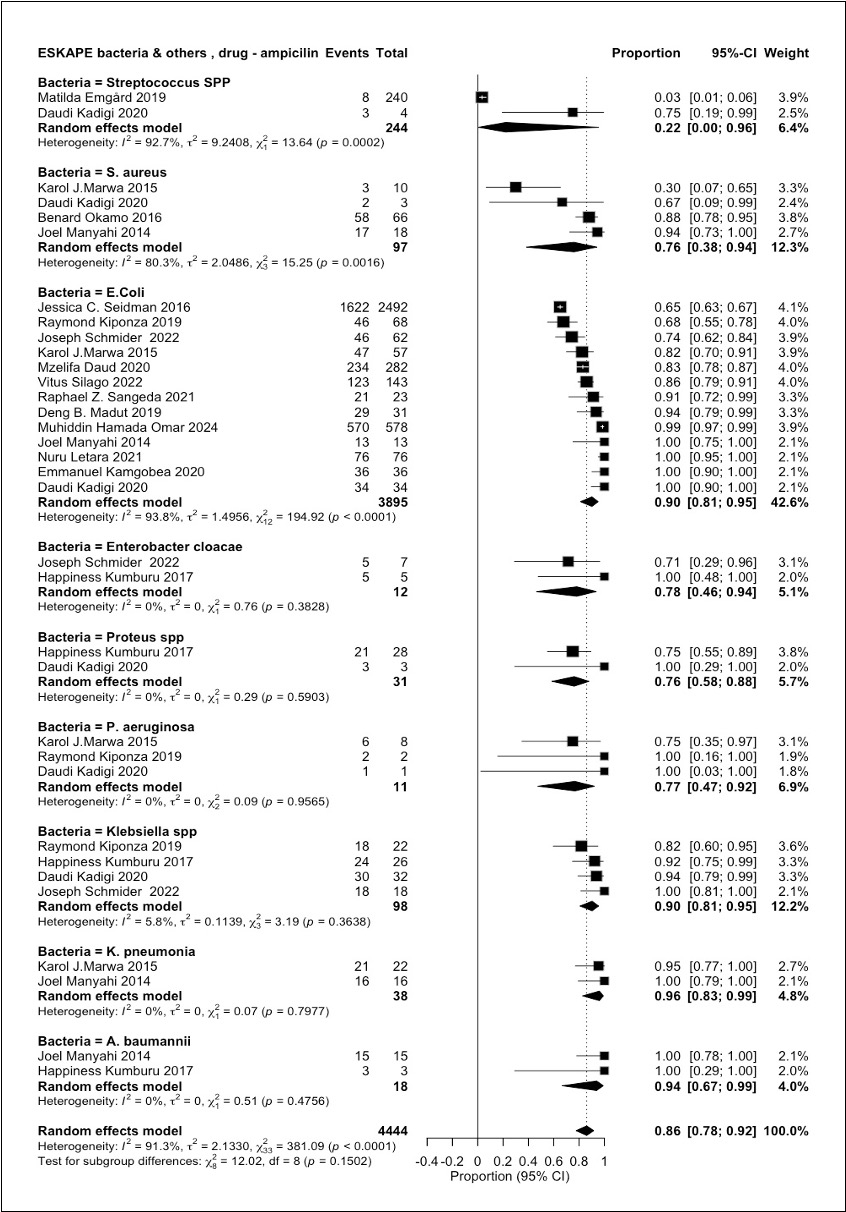

**S1 Fig: Ampicillin resistance patterns among various pathogens**

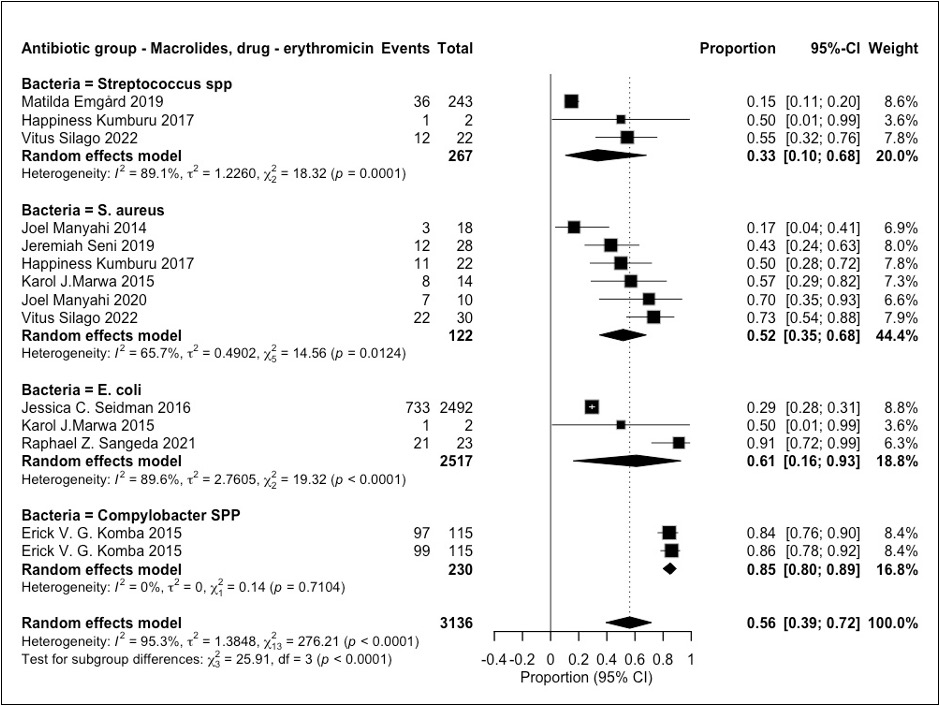

**S2 Fig: Erythromycin resistance patterns among various pathogens**

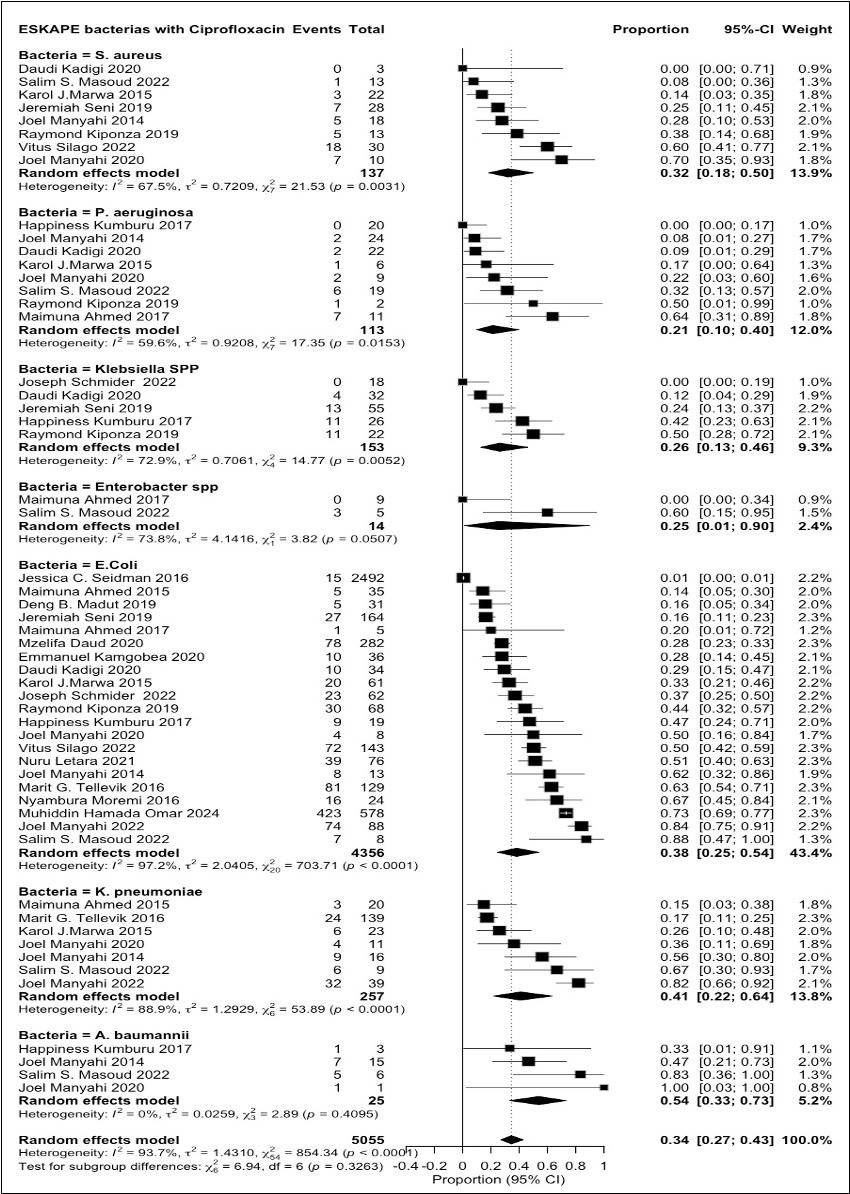

**S3 Fig. Ciprofloxacin resistance patterns among various pathogens**

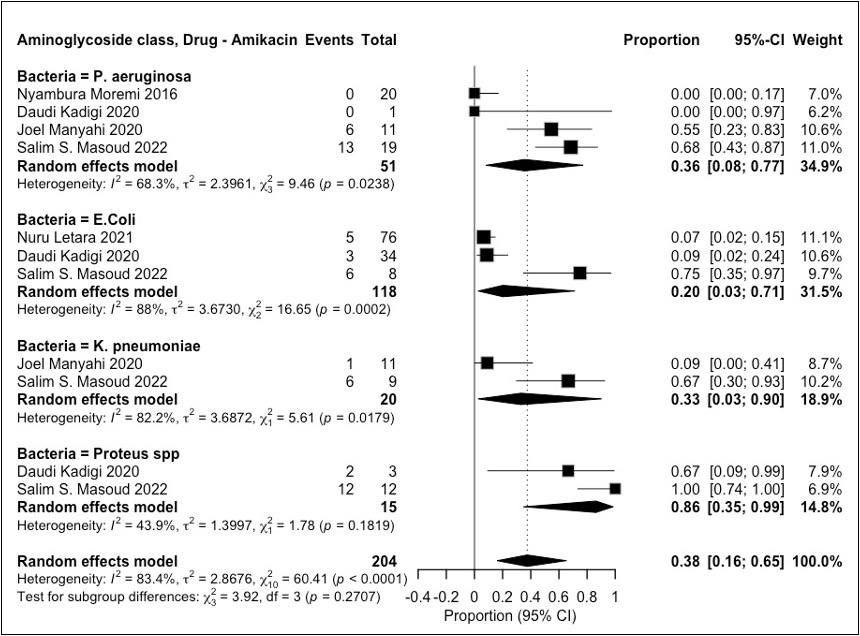

**S4 Fig. Amikacin resistance patterns among various pathogens**

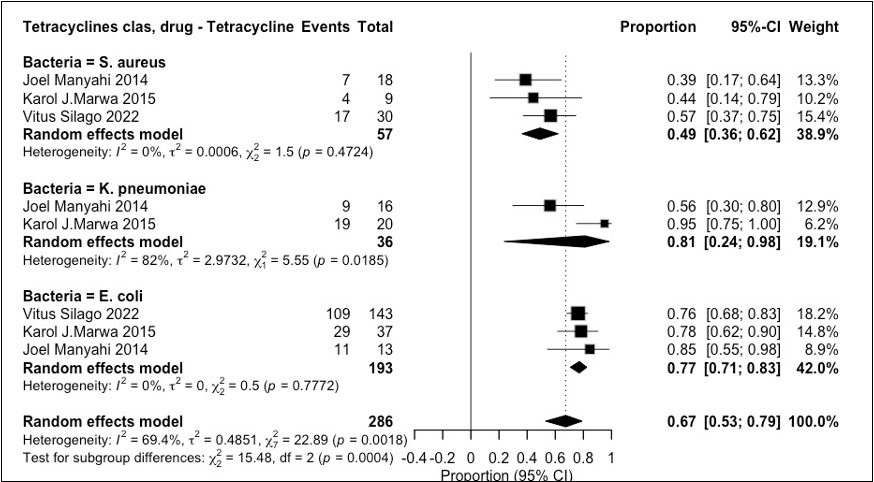

**S5 Fig. Tetracycline resistance patterns among various pathogens**

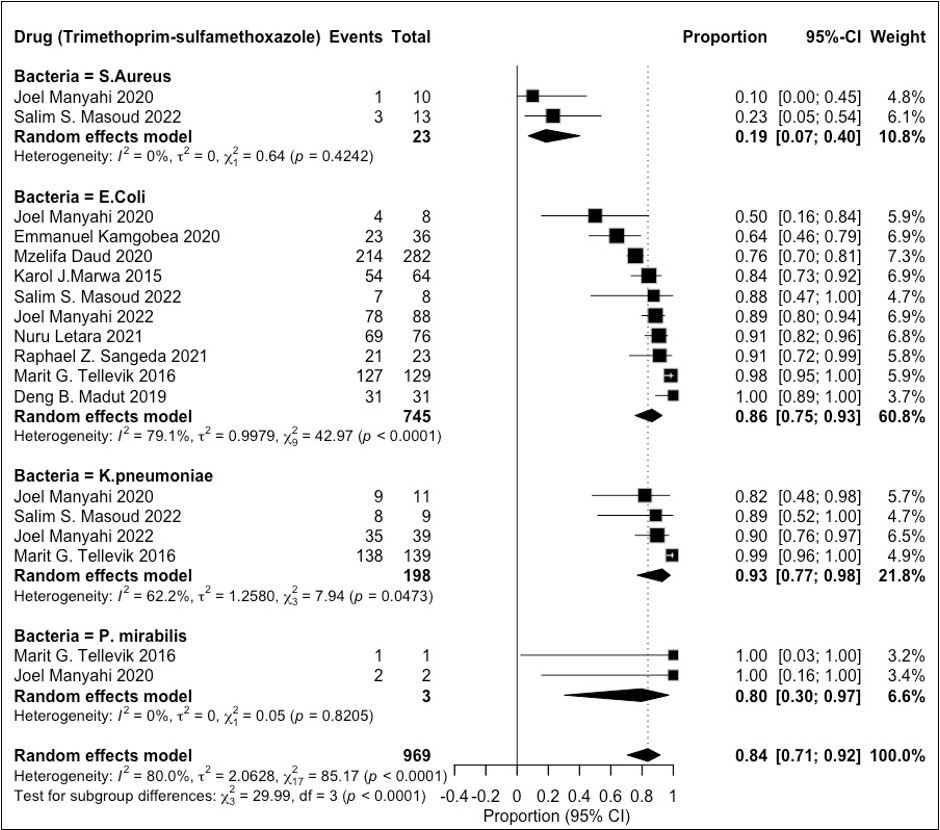

**S6 Fig. Trimethoprim-sulphamethoxazole resistance patterns among various pathogens**

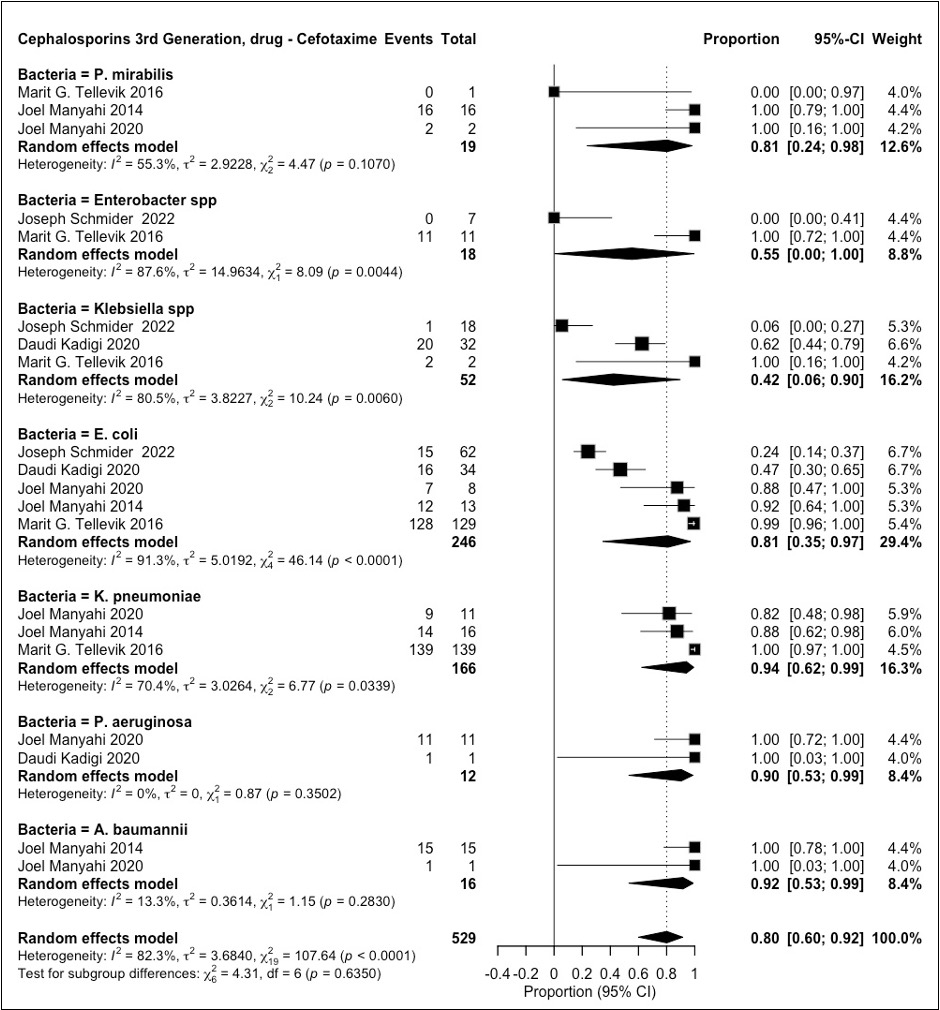

**S7 Fig. Cefotaxime resistance patterns among various pathogens**

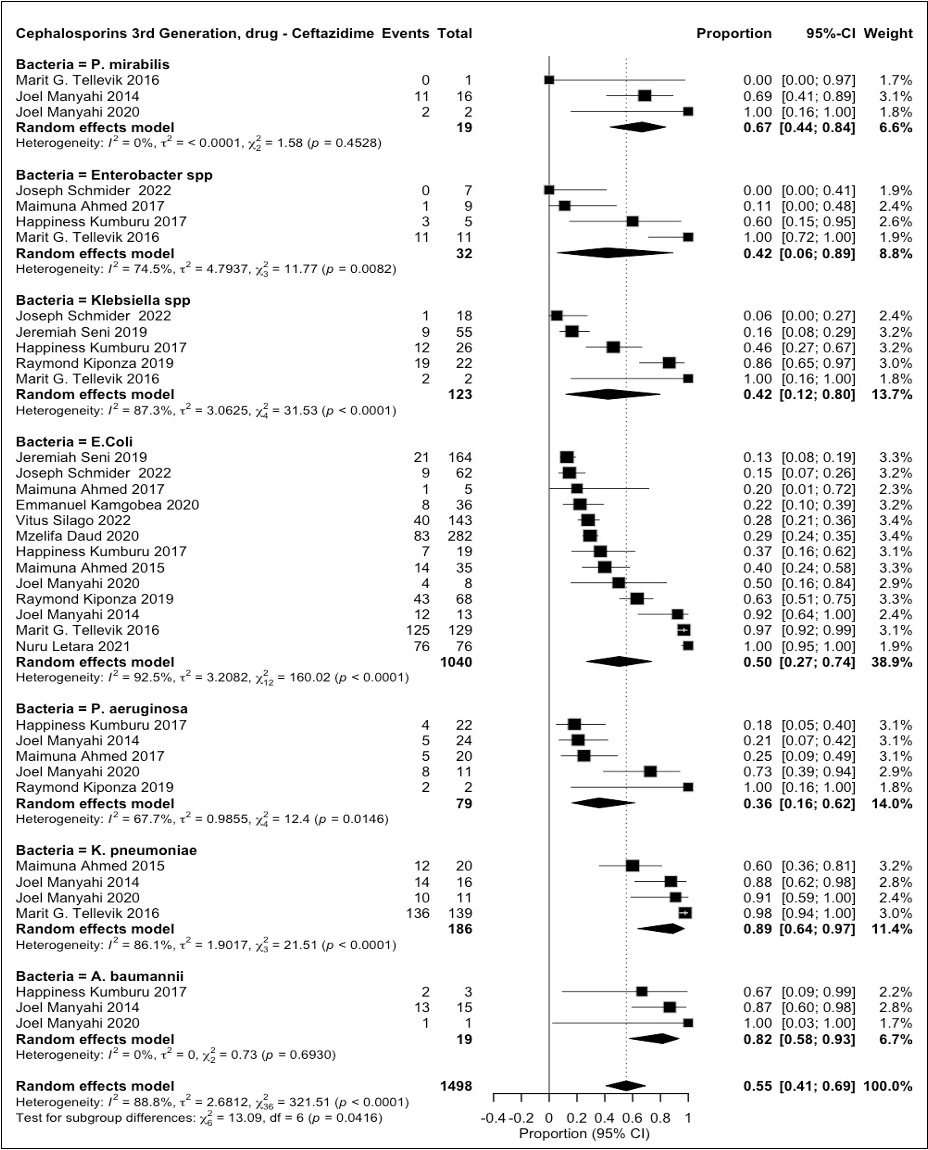

**S8 Fig. Ceftazidime resistance patterns among various pathogens**

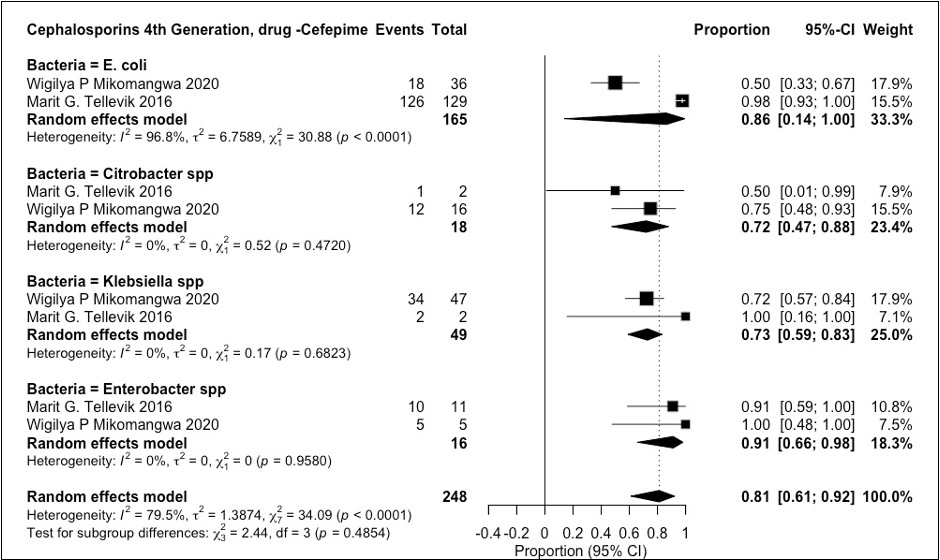

**S9 Fig. Cefepime resistance patterns among various pathogens**

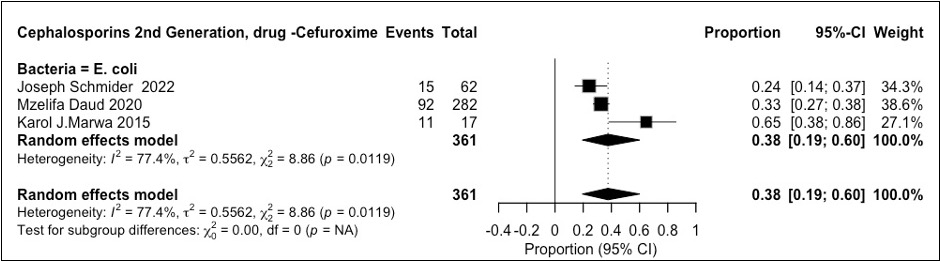

**S10 Fig. Cefuroxime resistance patterns with E.coli**

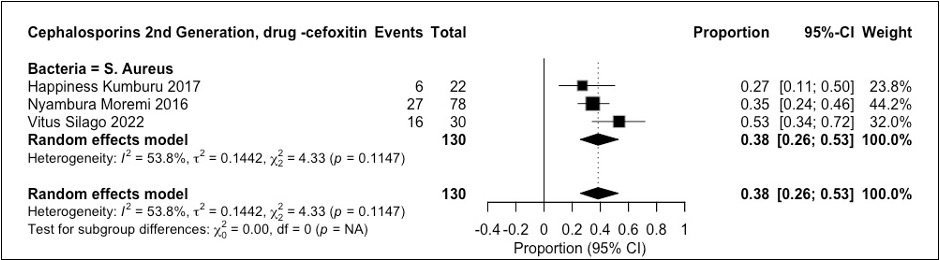

**S11 Fig. Cefoxitin resistance patterns among various pathogens**

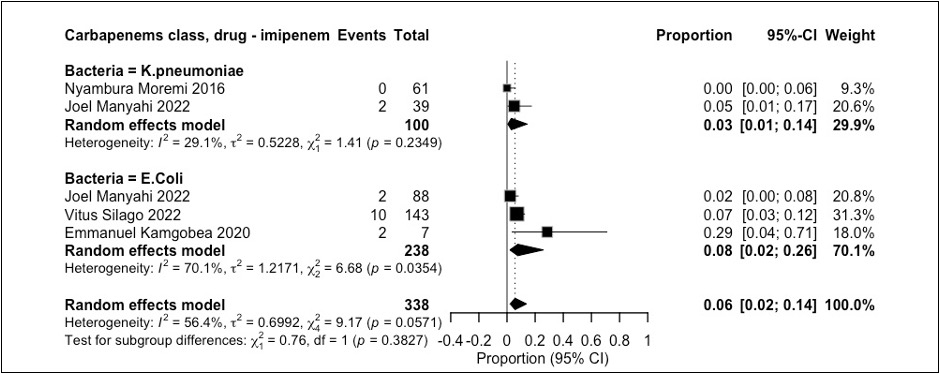

**S12 Fig. Imipenem resistance patterns among various pathogens**

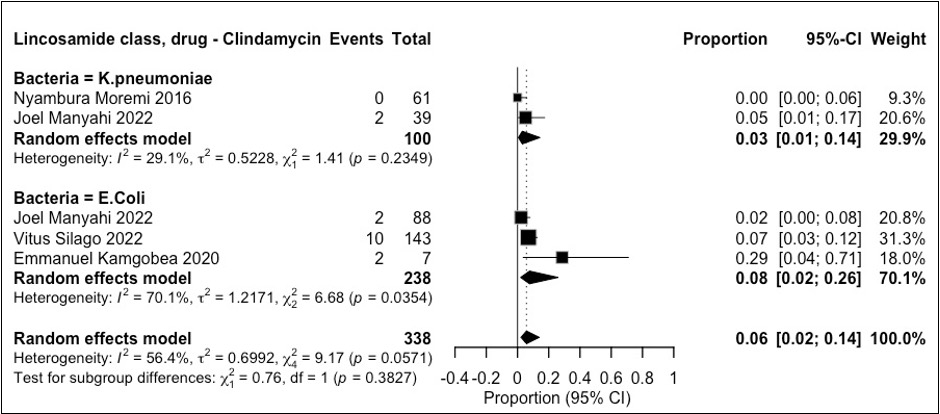

**S13 Fig. Clindamycin resistance patterns among various pathogens**

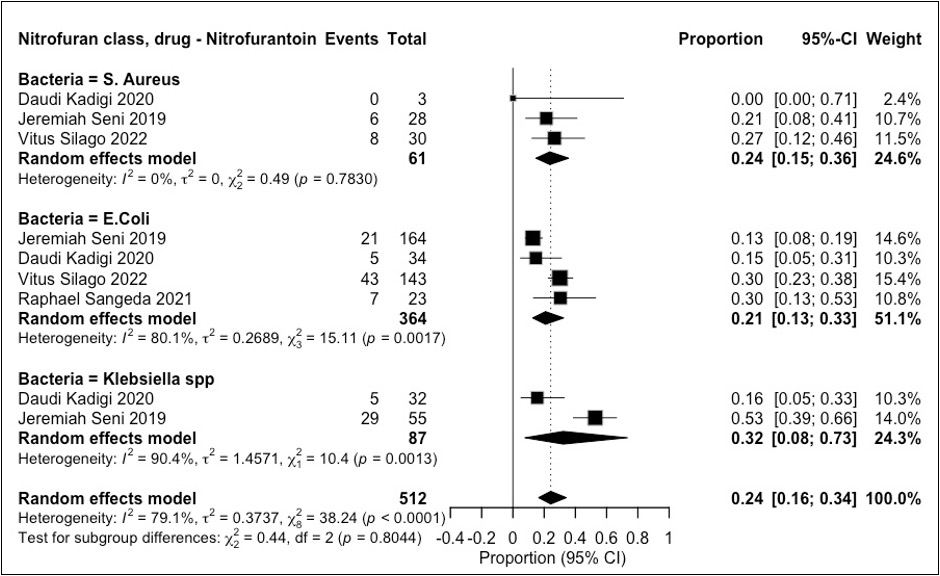

**S14 Fig**. **Nitrofurantoin** **resistance patterns among various pathogens**
